# Global patterns and drivers of influenza decline during the COVID-19 pandemic

**DOI:** 10.1101/2022.07.15.22277497

**Authors:** Francesco Bonacina, Pierre-Yves Boëlle, Vittoria Colizza, Olivier Lopez, Maud Thomas, Chiara Poletto

**Author notes:** Corresponding author: Chiara Poletto.

## Abstract

Influenza circulation declined during the COVID-19 pandemic. The timing and extent of decline and its association with interventions against COVID-19 were described for some regions. Here, we provide a global analysis of the influenza decline between March 2020 and September 2021 and investigate its potential drivers. We computed influenza change by country and trimester relative to the 2014-2019 period using the number of samples in the FluNet database. We used random forests to determine important predictors in a list of 20 covariates including demography, weather, pandemic preparedness, COVID-19 incidence, and COVID-19 pandemic response. With a regression tree we then classified observations according to these predictors. We found that influenza circulation decreased globally, with COVID-19 incidence and pandemic preparedness being the two most important predictors of this decrease. The regression tree showed interpretable groups of observations by country and trimester: Europe and North America clustered together in spring 2020, with limited influenza decline despite strong COVID-19 restrictions; in the period afterwards countries of temperate regions, with high pandemic preparedness, high COVID-19 incidence and stringent social restrictions grouped together having strong influenza decline. Conversely, countries in the tropics, with altogether low pandemic preparedness, low reported COVID-19 incidence and low strength of COVID-19 response showed low influenza decline overall. A final group singled out four “zero-Covid” countries, with the lowest residual influenza levels. The spatiotemporal decline of influenza during the COVID-19 pandemic was global, yet heterogeneous. The sociodemographic context and stage of the COVID-19 pandemic showed non-linear associations with this decline. Zero-Covid countries maintained the lowest levels of reduction with strict border controls and despite close-to-normal social activity. These results suggest that the resurgence of influenza could take equally diverse paths. It also emphasises the importance of influenza reseeding in driving countries’ seasonal influenza epidemics.

## Introduction

Starting with the global spread of SARS-CoV-2, observations of a sharp decline in influenza circulation were reported. In spring 2020, the flu season was shortened in some northern-hemisphere and tropical countries [1,2]. During the following 18 months, influenza incidence showed an all-time low in New Zealand [3], Australia [4], the United States [5–7] and the WHO European Region [8]. The circulation was still low in 2021.

The measures adopted in response to the COVID-19 pandemic are likely to have hindered influenza transmission at the same time, since the routes of transmission are identical. Indeed, influenza decline, as well as that of other transmissible diseases, coincided with non-pharmaceutical interventions against COVID-19 [2,6,7] [9,10].

Understanding how this decline occurred may help interpret the current influenza trends and anticipate future viral circulation. While the issue has been described for specific countries or regions [2–5,7,8,11–13], little work has been done at the global scale [14,15] [16].

Here we provide a global quantitative analysis of the influenza reduction based on the Global Influenza Surveillance and Response System FluNet database [16,17]. We considered the period between March 2020 and September 2021 and estimated influenza reduction by country and trimester relative to a pre-pandemic period (2014-2019). We identified geographical, demographical, health preparedness and COVID-19 status characteristics predictive of influenza decline using random forests and clustered observations with similar decline in time and space using a regression tree.

## Methods

### Overview of the methods

We used data from the FluNet influenza repository [16,17] to quantify the global influenza change during the COVID-19 pandemic (March 2020 to September 2021) compared to the pre-pandemic period (December 2014 to December 2019). We mapped influenza decline by country and trimester. We then used random forests to identify the most significant predictors of decline and a regression tree to classify countries-trimesters based on these predictors. Potential predictors included a wide range of covariates, among them country factors (geographical, meteorological, demographic and health preparedness factors) and variables associated with the COVID-19 pandemic assembled from sources detailed below.

### Influenza data and definition of influenza reduction

The FluNet influenza repository [16,17] provides weekly counts of influenza specimens by country. For our analysis we considered records from 2014 to 2021. To account for influenza seasonality, we defined 13 weeks-long “influenza trimesters” beginning on the first Monday following December 11, March 12, June 11 and September 11. These dates were chosen so that the middle of the December 11 trimester coincided with the peak of a typical influenza circulation in the northern hemisphere. As the trimesters correspond approximately to the astronomical seasons, we refer to these as winter, spring, summer and autumn respectively. Data from FluNet was aggregated by country and trimester. The 20 trimesters from winter 2014-15 (Dec 2014-Mar 2015) to autumn 2019 (Sep 2019-Dec 2019) defined the reference “pre-pandemic” period, the six trimesters from spring 2020 to summer 2021 the “pandemic” period.

The winter 2020 trimester (from Dec 2019 to Mar 2020) was excluded as it overlapped the period of COVID-19 emergence. We also discarded trimesters having less than 10 processed influenza specimens per week on average and those typically unaffected by influenza epidemics (i.e. having less than 5% of the annual positive cases on average during the pre-pandemic period, essentially “summer” in the northern hemisphere and “winter” in the southern hemisphere).

For the “pandemic” trimesters, we computed the percentage of influenza positive cases as the ratio of positive to positive plus negative samples during the trimester (adding 0.5 to avoid division by zero issues). We computed the “log relative influenza level” as the base-10 logarithm of the ratio between the percentage of positive cases during a trimester and the average percentage of positive cases in the corresponding pre-pandemic trimesters [7,13].

### Variables for prediction of influenza reduction

We collected the covariates described in Table 1 from public sources and IATA. Additional details on computation are provided in the supplementary material.

**Table 1.** Definition, computation and source of the variables used as predictors of influenza change.

| Variable | Description | Source | Min, max |
| --- | --- | --- | --- |
| age | Median age of the country population | [18], [19] | 15.1, 48.2 |
| longitude | Population-weighted average of longitude for cities with more than 300K inhabitants by country or country capital longitude, from -180 (W) to 180 (E) | [20] | -100.7, 174.4 |
| latitude | Population-weighted average of latitude for cities with more than 300K inhabitants by country or country capital latitude, from -90 (S) to 90 (N) | [20] | -38.7, 60.4 |
| T | Average temperature (in Celsius degrees) over the country and trimester | [21] | -8.8, 37.8 |
| RH | Average relative humidity over the country and trimester | [21] | 17.3, 93.5 |
| IDVI | Infectious Disease Vulnerability Index (IDVI), country level indicator of the vulnerability to health emergencies from 0 (most vulnerable) to 1 (less vulnerable) | [22] | 0.15, 1 |
| COVID-19 daily cases | Average daily reported cases of COVID-19 per million inhabitants | [23] | 0, 553.5 |
| workplace presence reduction | Median percentage of reduction of daily presence in workplaces. Reduction from the first 5 weeks in 2020 in the same location. | [24] | -22.5%, 69.0% |
| reduction of international flights | Average percentage of reduction in the inbound and outbound air passengers of the country for each trimester with respect to the same trimester of 2019 | [25] | -16.8%, 100% |
| nb days of school closure | Number of days over the trimester where policies related to schools and universities closure were implemented | [26] | 0, 91 |
| nb days of workplace closure | Number of days over the trimester where policies related to workplaces closure were implemented | [26] | 0, 91 |
| nb days of public event restrictions | Number of days over the trimester where policies related to event restrictions were implemented | [26] | 0, 91 |
| nb days of gathering restrictions | Number of days over the trimester where policies related to social gathering restrictions were implemented | [26] | 0, 91 |
| nb days of public transport restrictions | Number of days over the trimester where policies related to public transport restrictions were implemented | [26] | 0, 91 |
| nb days of stay at home requirements | Number of days with "shelter-in-place" and otherwise confine to the home orders | [26] | 0, 91 |
| nb days of international travel restrictions | Number of days with airport screening, quarantine of arrival passengers or restrictions of international travels | [26] | 0, 91 |
| nb days of facial covering requirements | Number of days with policies on the use of facial coverings outside the home | [26] | 0, 91 |
| nb days of testing implementation | Number of days with government policy on who has access to testing for current infection (PCR tests) | [26] | 0, 91 |
| nb days of contact tracing implementation | Number of days with government policy on contact tracing after a positive diagnosis | [26] | 0, 91 |
| nb days of elderly shielding | Number of days with policies to protect older adults (as defined locally) in long-term care facilities and/or community and home-based settings | [26] | 0, 91 |

### Clustering and regression tree analysis

We used the VSURF algorithm based on random forests (RFs) to select the covariates that were highly predictive of influenza reduction [27]. Importance is defined as the increase in prediction-error when the variable of interest is randomly reshuffled across observations. We discarded variables with close to zero importance in a univariable analysis. Then, we carried out a forward selection of predictors, including variables in their order of importance one at a time. Following Breiman’s rule [28], we retained the model with the least variables having prediction error less than the minimum prediction error plus one standard deviation.

Using the variables selected above, we fit a regression tree in order to obtain an interpretable model [28]. The details of the approach are provided in the supplementary material.

Analyses were performed with R version 4.2 and packages *vsurf* [27] and *rpart* [29].

### Robustness and sensitivity analyses

The details of the robustness checks and the sensitivity analysis are reported in the supplementary material. In summary, we checked the robustness of the regression analysis to stochastic fluctuations in the dataset and to criteria for including the FluNet records in the analysis; we explored alternative definitions for covariates: COVID-19 daily deaths instead of COVID-19 daily cases; Oxford COVID-19 Government Response Tracker stringency index instead of governmental response [26]; alternative Google mobility reports instead of presence in workplaces. We also explored separate inclusion of age and IDVI as these were highly correlated (*ρ*_*Spearman*_*=0*.*87, p*_*val*_*<0*.*01*).

## Results

### Decline of influenza in space and time

One hundred sixty-six (166) countries contributed data to FluNet between December 2014 and September 2021. Figure 1A shows the time course of the reports. In the pre-pandemic period, the percentage of positive tests varied seasonally between 4% and 33%, with major peaks during seasonal epidemics in northern countries and lower peaks for southern countries. The global number of tests for influenza remained within the range of historical levels throughout the whole COVID-19 pandemic period, but the percentage of influenza positive tests dropped sharply, to a minimum level of 0.04% during the months of July and August 2020.

**Figure 1.**
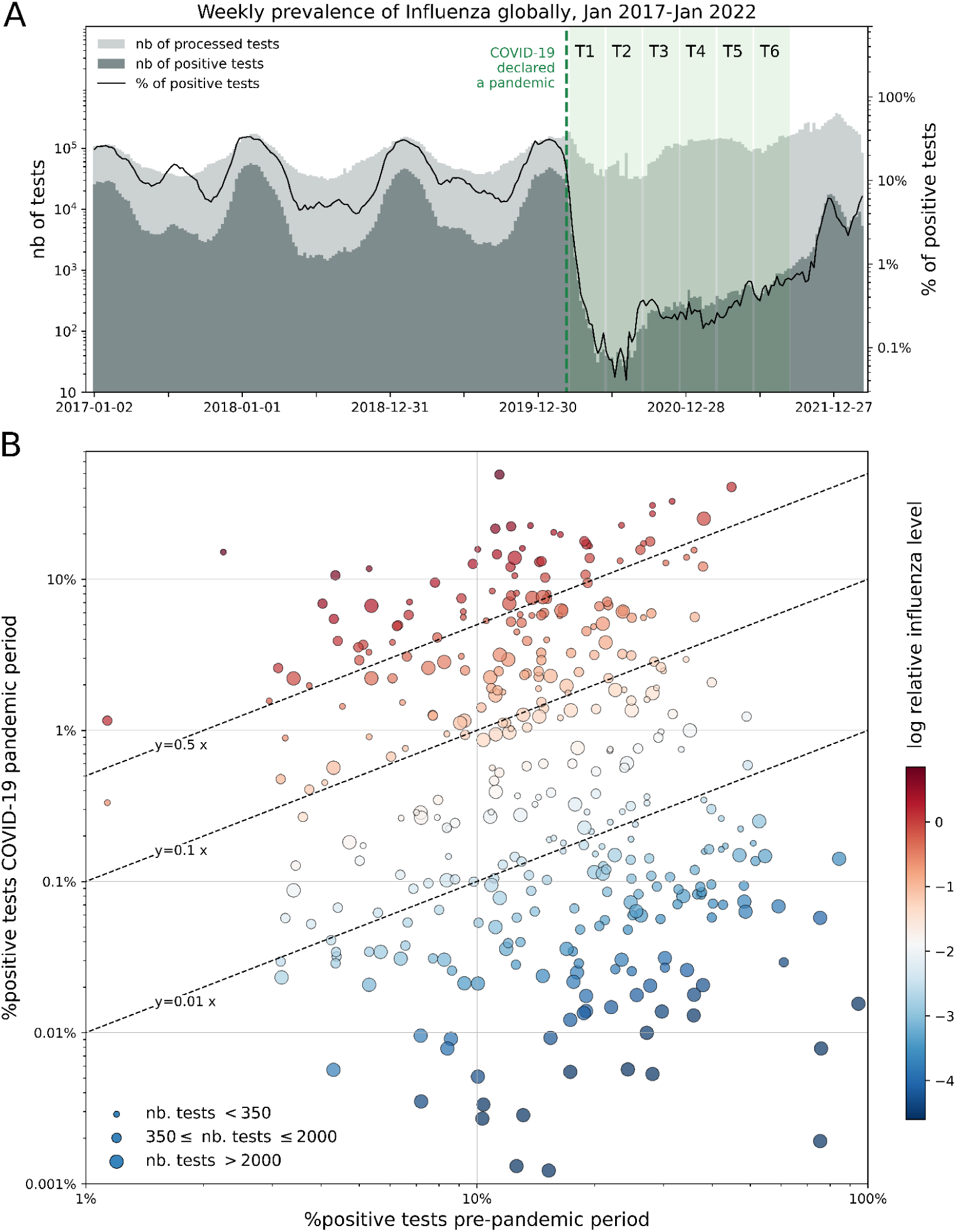
Change in influenza circulation during the COVID-19 pandemic relative to the pre-pandemic period. **A** Weekly counts of processed and positive tests of influenza reported to FluNet for all 166 countries included in the database from Jan 2017 to Jan 2022. The green shaded area indicates the COVID-19 pandemic period considered in the study. The six blocks indicate the trimesters. The week in which COVID-19 was declared a pandemic by WHO is reported as reference. **B** Percentage of positive tests for the pre-pandemic and COVID-19 pandemic periods (Dec 2014 - Dec 2019 and Mar 2020 - Sep 21, respectively), for all 376 countries and trimesters satisfying the filtering criteria on the FluNet data. For each country-trimester, the x coordinate is the average percentage of positive tests of the five years included in the pre-pandemic period. The size of the dots is proportional to the number of samples found in FluNet for the pandemic period. Dots’ colour indicates the log relative influenza level.

One hundred twelve countries remained for analysis during the pandemic, contributing 376 trimester-country observations (Table S1). The percentage of influenza positive tests varied across countries and trimesters over five orders of magnitude compared to only two orders of magnitude over the pre-pandemic period (Figure 1B). For 135 out of the 376 observations, the percentage of positive influenza tests was more than 100 times smaller than expected. The reduction of influenza positivity could be dramatic, as shown by the 0 positive tests out of 26114 processed tests reported in Japan during Spring 2021, compared to the average 75% expected in the pre-pandemic period. An increase in the percentage of positive tests was seen in 22 observations: This was for example the case for Haiti during Winter 2020-21, where the percentage of positive tests was 15% compared to an expected 2.2% before the pandemic.

The spatial variation of the influenza decline is mapped in Figure 2 over the 6 pandemic trimesters. For the majority of countries, the decline remained limited during Spring 2020, with 46 out of 65 countries reporting less than 90% reduction from the pre-pandemic period (i.e. log relative influenza level > -1). The decline became more pronounced in the subsequent trimesters, especially in North America, Europe, Mexico and Japan during the winter 2020-21 and spring 2021. The decline was strong also in the majority of Southern-hemisphere countries during both summer 2020 and summer 2021. Conversely, a number of countries in South Asia (e.g. Bangladesh, Afghanistan), Africa (e.g. Mali, Senegal, Nigeria, Kenya, Zambia) and Central America (e.g. Honduras, Haiti) showed limited influenza reduction throughout the whole COVID-19 pandemic period (log relative influenza levels > -1). The levels of reduction changed over the period. Interestingly, the log relative influenza level was as low as -2.4 during summer 2020 in China but increased again starting autumn 2020. A similar increasing trend was observed also in a few other countries, e.g. in Kenya and Nigeria.

**Figure 2:**
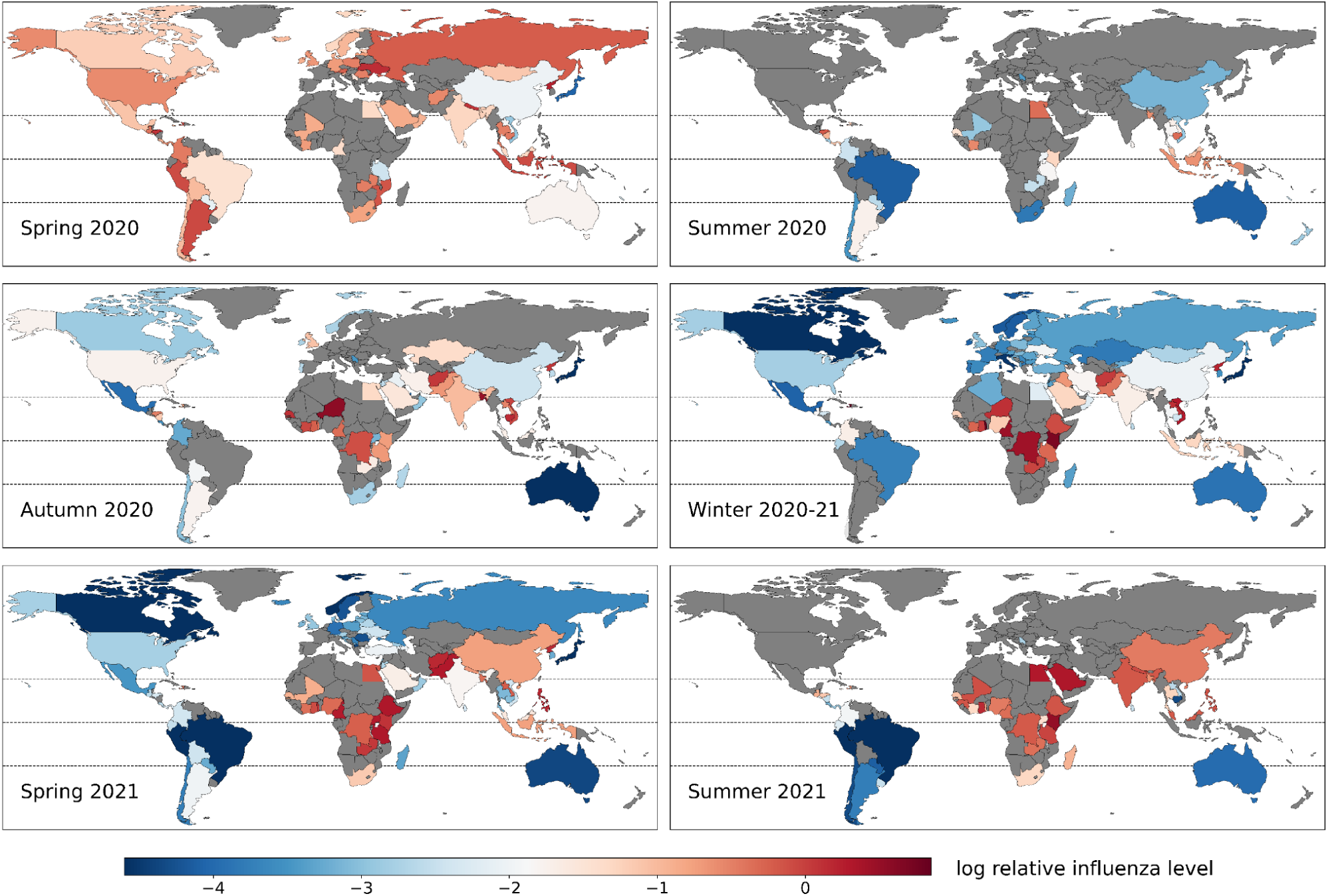
Influenza decline during the first 18 months of COVID-19 pandemic by countries and trimesters. Maps of the log relative influenza level for the 6 trimesters considered in the analysis. The grey colour indicates countries-trimesters not included in the analysis.

### Clustering and regression tree analysis

The analysis was carried out on 93 countries, totalling 330 country-trimester observations. Among the 20 covariates tested, 11 were selected as predictors of the log relative influenza level (Figure 3). Sociodemographic, preparedness, geographical, weather and COVID-19 management aspects contributed all to explaining the changes, though COVID-19 daily cases and IDVI were the most important.

**Figure 3:**
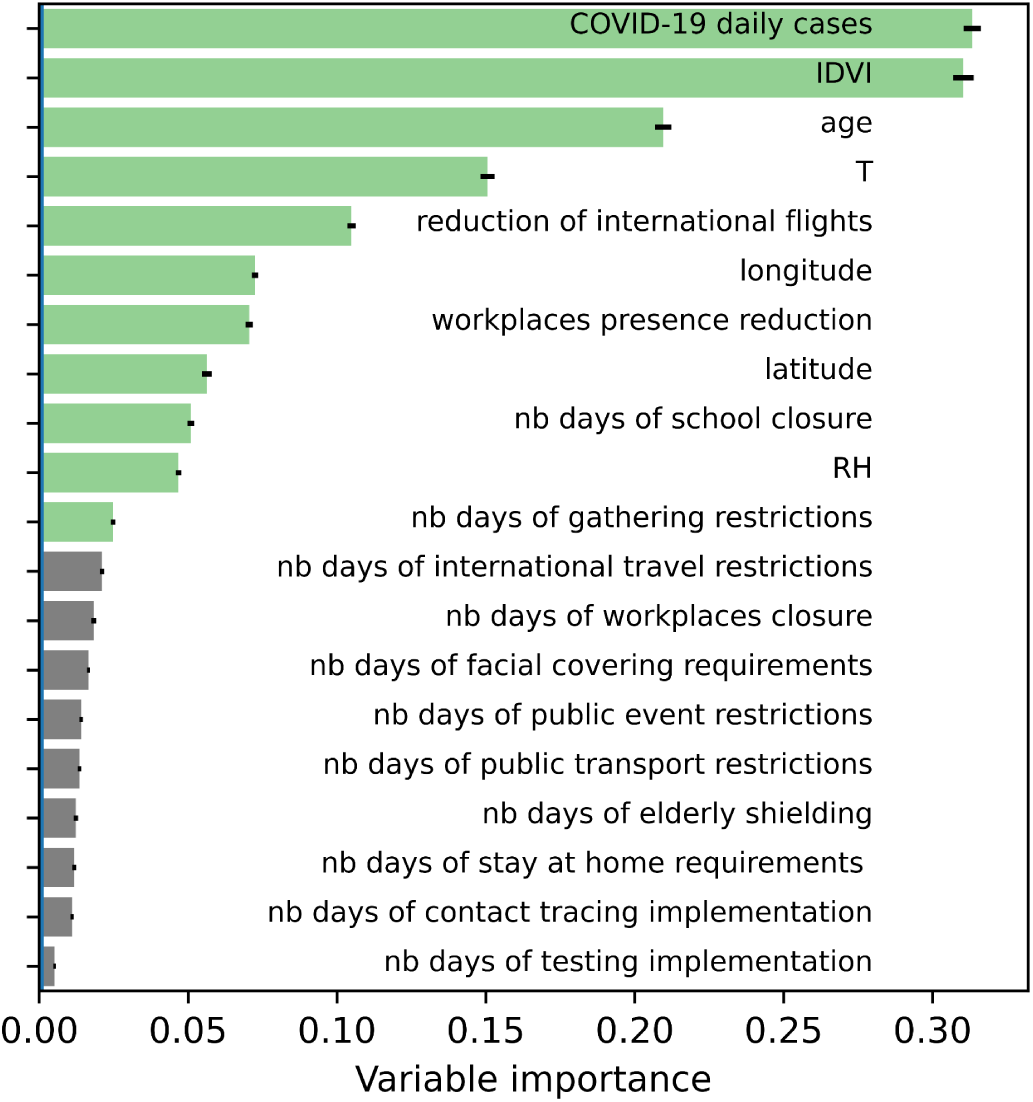
Importance of covariates predicting influenza decline in random forest analysis. Importance of covariates as predictors of the log relative influenza level. In green the 11 covariates selected as significant to build the model with the minimum prediction error following the Breiman’s rule. Black segments show the standard deviations of the importance.

The full regression tree built from the data accounted for 69% of the variance of the log relative influenza level (*R*^2^=0.69) (Figure S3, Table S2). To interpret the relationships between the selected variables and the country-trimesters, we focus here on the first four splits based on IDVI, COVID-19 daily cases, longitude, and workplace mobility reduction (Figure 4A). The five groups identified by these splits (labelled 1 to 5, Figure 4A) showed a gradient in average log relative influenza level ranging from -2.3 (reduction by 99.5%) to -0.7 (reduction by 80%). How the observations in each group rank with respect to the whole dataset is shown in Figure 4B. Group 1 included 109 countries-trimesters with high influenza decline, corresponding to the lower quartile of the whole dataset distribution. This group was characterised by high IDVI (median value corresponding to the 71th pc. of the whole dataset), high COVID-19 daily cases (83rd pc.), old population (70th pc.), low temperatures (25th pc.). Median reduction of workplace presence and median number of days with school closure were close to the whole-population median but were higher than other groups, except for group 4 discussed below. Population gathering restrictions were especially high (82nd pc.). The corresponding countries-trimesters included countries in Europe and North America during the 2020-21 influenza season, countries in temperate South America, and high-IDVI countries in Central America and Tropical Asia (Table S2 in the supplementary material).

**Figure 4:**
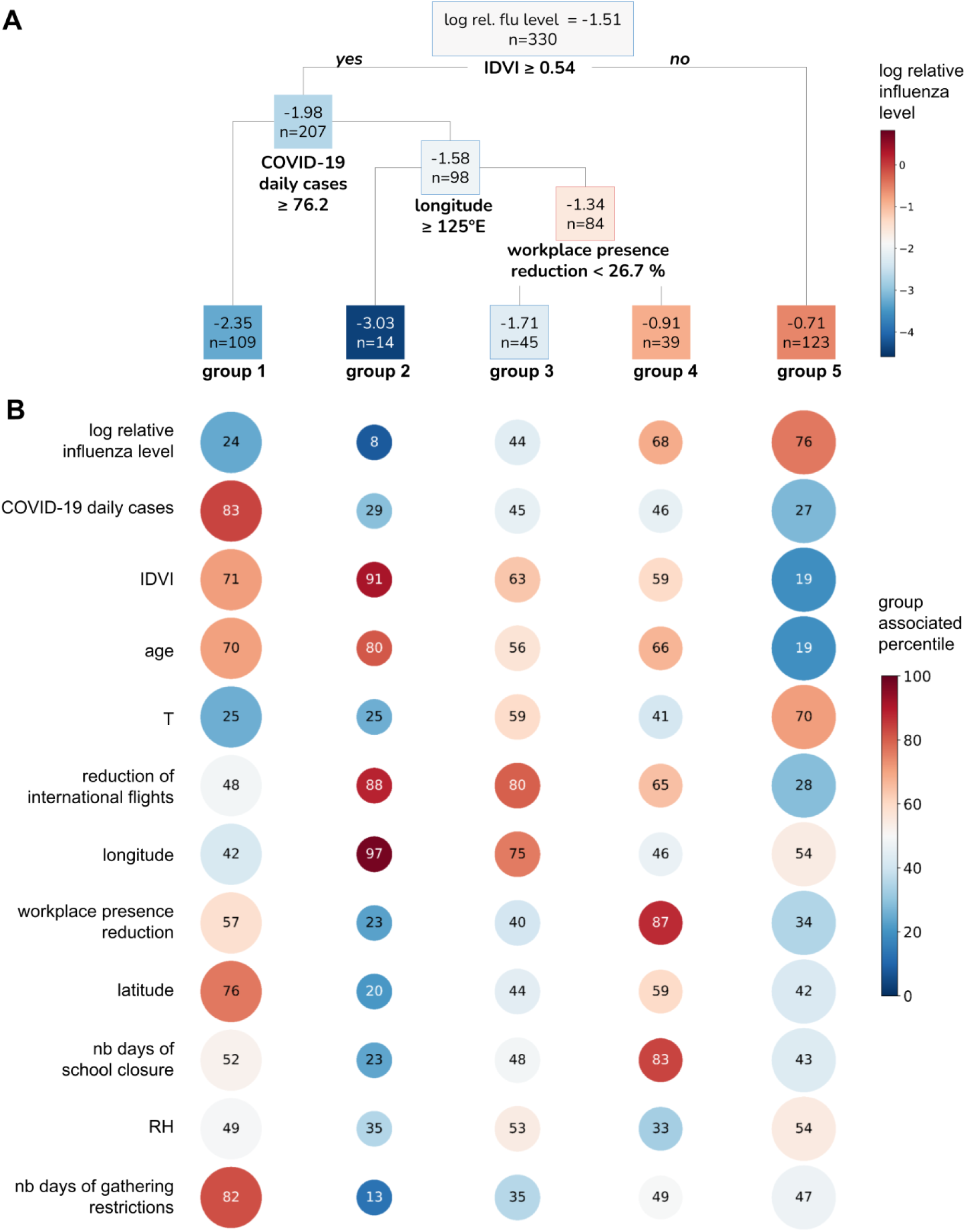
Regression tree analysis of influenza decline and characteristics of the identified subgroups. **A** Regression tree obtained with the variables selected in Figure 3. We report here the first four splits, which partition the observations in five groups. The full tree is reported in Figure S4 of the supplementary material. For each node the average log relative influenza level and the number of observations are reported (the former is also indicated with a colour scale). **B**. Characteristics of each group. For each variable the colour of the circle indicates the percentile of the whole dataset distribution the median of the group corresponds to. The percentile value is also indicated inside the circle. The size of the circle increases with the number of observations of the group.

Group 2 was the smallest and clustered observations with the largest influenza decline (median log relative influenza level corresponding to the least 8% of all data points). It gathered all observations from Australia, Japan, New Zealand and South Korea. These country-trimesters showed low COVID-19 daily cases (29th pc.), high IDVI (91st pc) and high reduction of international flights (88th pc.). Reduction of workplace presence, and number of days of school closure and gathering restrictions were comparatively low (23rd, 23rd, 13rd, pcs., respectively).

Group 3 corresponded to 45 observations with intermediary log relative influenza level. Covariates were also close to the median of all data points. Singapore from autumn 2020 to autumn 2021 is part of this group (larger tree in Figure S4). Covariates of these observations are close to the second group - e.g. high influenza reduction, low COVID-19 daily cases, high reduction of international flights. Other observations of group 3 (e.g. Southeast Asia countries, such as Malaysia, Vietnam, Indonesia and Thailand) were similar to Singapore, but had lower population’s age and IDVI. They showed, however, a more limited influenza decline.

Group 4 had 39 observations corresponding to Europe and North America during the spring 2020 trimester. At this period, influenza decline was limited (median log relative influenza level corresponding to the 68th pc. of all data points), but there were already a strong response to the COVID-19 pandemic as quantified e.g. by the reduction in the workplace presence (87th pc.) and number of days of school closure (83rd pc.).

Finally, group 5 included 123 country-trimesters with the lowest decrease in influenza relative to the pre-pandemic period (log relative influenza level 76th pc.). In this group, there was a low number of COVID-19 cases (27th pc.), young population (19th pc.), low IDVI (19th pc.) and high temperatures (70th pc.). The response to the COVID-19 pandemic was mild, with limited reduction of international flights (28th pc.), as well as workplace presence reduction (34th pc.) and number of days of school closure (43rd pc.) small compared to the whole population. This group was largely formed by tropical countries, e.g. in Africa, South and Southeast Asia, Central America and the Caribbean (Table S2 of the supplementary material).

### Robustness and sensitivity analyses

Variable selection and tree structure were robust to stochastic fluctuations. The five-group classification was robust to small perturbations in the data set, as was the selection of predictive variables. In some cases, for example with different inclusion criteria for FluNet data, observations in Group 1 and Group 2 tended to cluster together. More details are reported in the supplementary material.

### Discussion

The systematic analysis of influenza circulation across all continents and climatic regions shows that the influenza decline was global during the spread of the COVID-19 pandemic. This decrease was heterogeneous across countries and trimesters between March 2020 and September 2021. Demographic, socio-economic, weather and COVID-19 characteristics explained a large part of this heterogeneity.

Influenza circulation is characterised by marked seasonal epidemics in temperate countries but a more complex annual pattern in the tropics [30]. Surveillance may be reinforced in epidemic times. Using the log relative influenza level allowed adjusting for such changes. We found that influenza declined nearly everywhere and remained low compared to the pre-pandemic period during the 18 first months of the COVID-19 pandemic. Importantly, the global number of influenza tests remained roughly the same in the pre-pandemic and pandemic period, ruling out change in surveillance as the likely explanation. The largest reduction was in summer 2020, and a progressive increase was seen again till September 2021. Temperate countries had the largest reduction, while it was limited in the tropics [11,14,31,32].

Influenza circulation could *a priori* change during the COVID-19 pandemic because of governmental measures, self-adopted behavioural changes and direct interaction with SARS-CoV-2. We indeed found that reduction of international flights, presence at workplaces, school attendance and mass gatherings explained part of the reduction, although the impact was non-linear. Initial strong restrictions against COVID-19 had to be relaxed in some low-resource countries [11,33,34] allowing renewed influenza circulation. Conversely, countries where a strong response against the COVID-19 pandemic could be maintained saw little influenza circulation, except in spring 2020 where strong local restrictions in Europe and the USA [5] likely occurred after the end of the influenza season. For the rest of the time, temperate countries in Europe, North America and South America that adopted a COVID-19 response centred over local restrictions by reducing workplace presence, school attendance and gatherings had large reduction in influenza circulation, irrespective of the reduction of international flights. This was very different in four “zero-Covid” nations (Australia, New Zealand, Japan and South Korea) where influenza dropped though local restrictions were limited [35], suggesting a key role for border controls in preventing seeding from abroad. Reducing international flights by 94-97% however did not prevent influenza introduction in Vietnam from neighbouring Cambodia [11] likely due to the difficulty of controlling land borders.

Limitation of gatherings or public events, imposed international travel restrictions and school closure were previously found to be the main drivers in suppressing influenza [12,13]. Actual behaviour, i.e. volume of flights rather than imposed international travel restrictions; or percentage presence at the workplace rather than mandatory reduction was however more predictive of influenza reduction than governmental restrictions. Behavioural proxies may indeed capture adhesion to restrictions that depended on place and stage of the pandemic [36–38].

Reduction of influenza could also stem from direct viral interference with SARS-CoV-2, for example through competition for cellular resources or interferon production [39,40]. Infection rates with influenza reportedly changed according to SARS-CoV2 status and vice versa [40]. In this respect, we found high influenza decline with high COVID-19 incidence in group 1, and low influenza decline with low COVID-19 incidence in group 5, but also low levels for both in zero-COVID countries. Under-reporting of COVID-19 cases may be an alternative explanation to low COVID-19 reporting in the low-income countries of group 5 [33,34,41].

The characterisation of influenza decline in space and time may come of use to analyse its resurgence over time. Loss of exposure to the influenza virus may lead to more severe waves or out of season waves [6,7] and may increase the susceptible pool, especially in children.

Already, influenza circulation was typically late in Europe as of spring 2022 [42]. Other epidemiological changes could occur regarding the exposed population and the seeding from the tropics [30] as global air transportation resumes. Deciding on the composition of the vaccine may also prove more difficult due to the change in the evolutionary dynamics of circulating strains [31].

Our study is affected by limitations. We assumed that influenza surveillance was not substantially altered during the pandemic period. The number of samples in the FluNet databases indeed did not change substantially over time, as many countries maintained influenza surveillance or quickly resumed it after initial disruption [3,4]. Influenza positivity rate may have been affected by changes in surveillance protocols due to the COVID-19 pandemic. We did not account for influenza vaccination, due to limited information at the global scale.

Vaccination rates are highly heterogeneous among countries [43]. While targeted recommendations increased coverage in the elderly during the last 2 seasons in 9 northern hemisphere countries and Australia [43], the efficacy of the influenza vaccine during the study period remains unknown. Those circulating in Southeast Asia during autumn 2020 were not included in the recommendations for the 2020–21 Northern Hemisphere season [11]. Last, we relied on the FluNet database, which integrates worldwide influenza records aggregating countries with highly diverse influenza surveillance quality and coverage. Results from the sensitivity analysis showed that the reported results were similar in varying exclusion criteria.

## Data Availability

Proprietary airline data are commercially available from the International Air Transport Association (https://www.iata.org/) database. Restrictions apply to the availability of these data, which were used under license, and so are not publicly available. All other data are openly available online at the references cited in the manuscript.

https://docs.google.com/spreadsheets/d/1PirC3iJ_yrlw9CoNL0_dkmXEI7DVVoJp/edit?usp=sharing&ouid=112424629266716674645&rtpof=true&sd=true

## Acknowledgements

We acknowledge financial support from the Municipality of Paris (https://www.paris.fr/) through the programme Emergence(s) to FB and CP; EU H2020 grants MOOD (H2020-874850) to PYB, VC and CP; Institut des Sciences du Calcul et de la Donnée.

## Data sharing

All data are available online from the references cited in the manuscript, except for airline data that were used under licence from the International Air Transport Association (https://www.iata.org/) and are not available for redistribution.

