## Supplementary material for "Global patterns and drivers of influenza decline during the COVID-19 pandemic"

**Additional methods** **2**
Definition of trimesters 2
Details on the computation of the log relative influenza level 2
Definition of covariates 2
Algorithms for the regression analysis 7
**Additional results** **8**
Countries included in the analysis 8
Regression tree 8
**Robustness checks and sensitivity analyses** **14**
Robustness of the variable selection procedure 14
Robustness of the tree structure optimization 14
Robustness of the tree under small perturbations of the dataset 14
Sensitivity of the variable selection under changes on the assumption made 14
**Bibliography** **16**

### Additional methods

#### Definition of trimesters

The trimesters considered in the analyses are periods of 13 weeks, defined so that the winter trimester best covers the typical period of flu epidemics in northern countries. Northern countries are defined as countries with latitude above the Tropic of Cancer. The winter trimester is identified based on FluNet data from 1995 to 2019, as follows. For each of the 365 possible starting dates of the trimester (a window of 91 days), we computed the annual proportion of positive cases falling in the period, averaged over all northern countries and years. We then define the winter trimester as the one that contains the highest annual proportion of positive cases. The spring, summer and autumn trimesters are identified accordingly. Also, we verified that the summer trimester according to this definition roughly contains the highest proportion of positive cases for southern countries. The winter, spring, summer and autumn trimesters obtained begin the first Monday following 12 December, 12 March, 11 June and 11 September, respectively. Certain years have 53 weeks instead of 52, thus trimesters may occasionally have 14 weeks.

#### Details on the computation of the log relative influenza level

Data reported on FluNet were partial or not consistent in some cases. Number of processed tests was sometimes different from the sum of positive and negative tests. In this case, the sum of positive and negative tests was used as the number of processed tests. When one of the three records (processed, positive and negative tests) was missing, this could be computed from the other two. The number of processed tests, when missing, was computed from the sum of positive and negative tests, the number of positive tests, when missing, was computed from the difference between processed and negative tests (provided the former was larger or equal to the later), and so on. We discarded weeks in which only processed and either positive or negative tests were present and the number of processed tests was smaller than the number of positive/negative tests. We also discarded weeks with only one record. Russia showed some irregularities with certain weeks having the number of processed tests nearly equal to the number of positive tests differently from the preceding or following weeks, signalling sudden changes in the data collection and sharing protocol. These weeks were removed from the analysis.

Before calculating the percentage of positive influenza tests, 0.5 positive cases are added to each country-trimester, so that the positivity rate always results greater than zero. This allows distinguishing countries without influenza and with a massive surveillance system from countries without influenza but processing only a few tests.

When working with percentages - e.g. the percentage of influenza positive samples or the percentage of annual influenza samples falling in a certain trimester - the centre of the distribution was computed from the closure of the geometric mean, that was proved to be a BLU (best linear unbiased) estimator, unlike the standard arithmetic mean [1].

#### Definition of covariates

<u>Definition of the covariates included in the main analyses:</u>

- **age**: median age of population, UN projection for 2020.

- **longitude**: longitude of the centre of population of the country in degrees, from -180 (W) to 180 (E). Longitude of the country is computed as the average longitude of all the cities of the country with more than 300K inhabitants. The average is weighted for the population size of each city. If there are no cities in the country with at least 300K inhabitants, the longitude of the capital is considered.
- **latitude**: latitude of the centre of population of the country in degrees, from -90 (S) to 90 (N). The latitude is calculated analogously to *longitude*.
- **T**: average temperature (in Celsius degrees) of the country-trimester. For each country, the temperature is computed as the average temperature of all the cities within the country with more than 300K inhabitants, weighted by the population size. If there are no cities in the country with at least 300K inhabitants, the capital is considered. Temperature data are taken from the ERA5 dataset, which provides hourly estimates of weather variables for all locations identified by a regular lat-lon grid of 0.25 degrees. The temperature of a city is calculated by looking at the closest grid point to the city and averaging the temperatures for the hours 0h00, 6h00, 12h00 and 18h00 of each day of the trimester.
- **RH**: average relative humidity, computed analogously to the *temperature*.
- **IDVI**: Score for the preparedness of a country in facing infectious diseases, from 0 (most vulnerable) to 1 (less vulnerable).
- **COVID-19 daily cases**: number of reported daily cases of COVID-19 per million of inhabitants averaged over the trimester.
- **workplace presence reduction**: median over the trimester of the daily percentage reduction of presence at workplaces.
- **reduction of international flights**: average percentage of reduction in the inbound and outbound air passengers of the country for each trimester with respect to the same trimester of 2019. The reduction for a trimester is calculated as the weighted average of the monthly reduction for the 4 months covering the trimester, with first and last months of the trimester, partially covered by the trimester, weighted 0.5, while the other months, fully covered by the trimester, weighted 1. The reduction for the month $m$ and year $y = 2020, 2021$ is defined as $1 - w_{m,y} / w_{m,2019}$, with $w$ being the number of passengers flying to or from the country.
- **nb days of school closure**: number of days over the trimester when policies related to schools and universities closure were implemented. The OxGRT dataset provides 2 daily variables: (i) the level of severity of the policy as measured on an ordinal scale (0=no measure, 1=altered openings for schools, 2=closing certain levels/categories of schools, 3=complete closure), and (ii) the geographical scope, i.e. whether that policy is enforced locally or nationally. Based on the values of these variables different definitions of school closure are possible - severity equal or above 1, 2, or 3, and each of these severity levels being implemented either locally or nationally. To choose the most convenient definition we used an unsupervised approach. We first computed the number of days with school closure for each data point (country-trimester) for all possible definitions. We then computed the distribution of the number of days with school closure over all data points and picked the definition with maximum resolution power, i.e. that maximises the number of observations with values not falling in the extremes. We obtained that schools are considered to be closed if certain levels/categories of schools were closed on a national scale or if there was at least one complete closure on a local scale.
- **nb days of workplace closure**: The severity levels defined in the OxGRT dataset were: 0=no measures, 1=recommend closings, 2=require closing for some sectors/categories of workers, 3=require closing for all-but-essential workplaces. With the unsupervised procedure described for school closure we obtained that

workplaces were defined as closed for stringency level at least 2 nationwide, or for stringency level 3 locally.

- **nb days of public event restrictions**: The severity levels defined in the OxGRT dataset were: 0=no measures, 1=recommend cancelling, 2=require cancelling. With the unsupervised procedure we obtained that public events were defined as closed when there was a countrywide enforcement.
- **nb days of gathering restrictions**: The severity levels defined in the OxGRT dataset were: 0=no restrictions, 1=restrictions above 1000 people, 2=restrictions between 101-1000 people, 3=restrictions between 11-100 people, 4=restrictions on gatherings of 10 people or less. With the unsupervised procedure we defined as gatherings restriction a nationwide ban of gatherings of more than 100 people.
- **nb days of public transport restrictions**: The severity levels defined in the OxGRT dataset were: 0=no measures, 1=recommend closing, 2=require closing. With the unsupervised procedure we obtained that public transports were defined as closed when a recommendation (level 1) was issued at local or national level.
- **nb days of stay at home requirements**: The severity levels defined in the OxGRT dataset were: 0=no measures, 1=recommend not leaving house, 2= require not leaving house with exceptions for 'essential' trips, 3=require not leaving house with minimal exceptions. With the unsupervised procedure described for school closure we obtained that staying at home was implemented for severity level 1 or more, locally or nationally.
- **nb days of international travel restrictions**: The severity levels defined in the OxGRT dataset were: 0=no restrictions, 1=screening arrivals, 2=quarantine arrivals from some or all regions, 3=ban arrivals from some regions, 4=ban on all regions or total border closure. With the unsupervised procedure we obtained that international travels were defined as enacted for severity level 3 or 4.
- **nb days of facial covering requirements**: The severity levels defined in the OxGRT dataset were: 0=no policy, 1=recommended, 2=required in some specified shared/public spaces with other people present, 3=required in all shared/public spaces with other people present, 4=required at all times regardless of location or presence of other people. With the unsupervised procedure we obtained that mask use was implemented for severity level 3 or 4.
- **nb days of testing implementation**: The severity levels defined in the OxGRT dataset were: 0=no testing policy, 1=only those who both (a) have symptoms AND (b) meet specific criteria, 2=testing of anyone showing COVID-19 symptoms, 3=open public testing. With the unsupervised procedure we obtained that testing policies were defined as implemented for stringency level 3.
- **nb days of contact tracing implementation**: The severity levels defined in the OxGRT dataset were: 0=no contact tracing, 1=not for all cases, 2=contact tracing for all identified cases. With the unsupervised procedure we obtained that contact tracing was defined as implemented for severity level 2.
- **nb days of elderly shielding**: The severity levels defined in the OxGRT dataset were: 0=no measures, 1=recommended isolation, hygiene, and visitor restriction measures in LTCFs and/or elderly people to stay at home, 2=narrow restrictions for isolation, hygiene in LTCFs, some limitations on external visitors and/or restrictions protecting elderly people at home, 3=extensive restrictions for isolation and hygiene in LTCFs, all non-essential external visitors prohibited, and/or all elderly people required to stay at home and not leave the home with minimal exceptions, and receive no external visitors. With the unsupervised procedure described for school closure we obtained that protection of elderly people was defined as implemented when it was enforced at least at level 2 locally or nationally.

<u>Definition of the covariates included in the sensitivity analyses:</u>

- **COVID-19 daily deaths**: number of reported daily deaths of COVID-19 per million of inhabitants averaged over the trimester.

- **station presence reduction**: median over the trimester of the daily percentage reduction of presence in public transport stations and transportation hubs.
- **recreation place presence reduction**: median over the trimester of the daily percentage reduction of presence at restaurants, bars, shopping malls and other recreation places.
- **home presence rise**: median over the trimester of the daily percentage rise of presence in residential places.
- **stringency index**: average of the daily stringency index provided by OxCGRT. This index combines eight indicators of containment and closure policies and an indicator regarding the presence of public information campaigns related to the pandemic. The daily index ranges from 0 for countries with no measures, to 100 for countries adopting maximally stringent policies regarding all nine indicators. Seven of the eleven indicators considered in the previous covariates are included in this index.

Covariate distributions: We provide in Figure S1 the distributions of the log relative influenza level and the covariates across countries-trimesters.

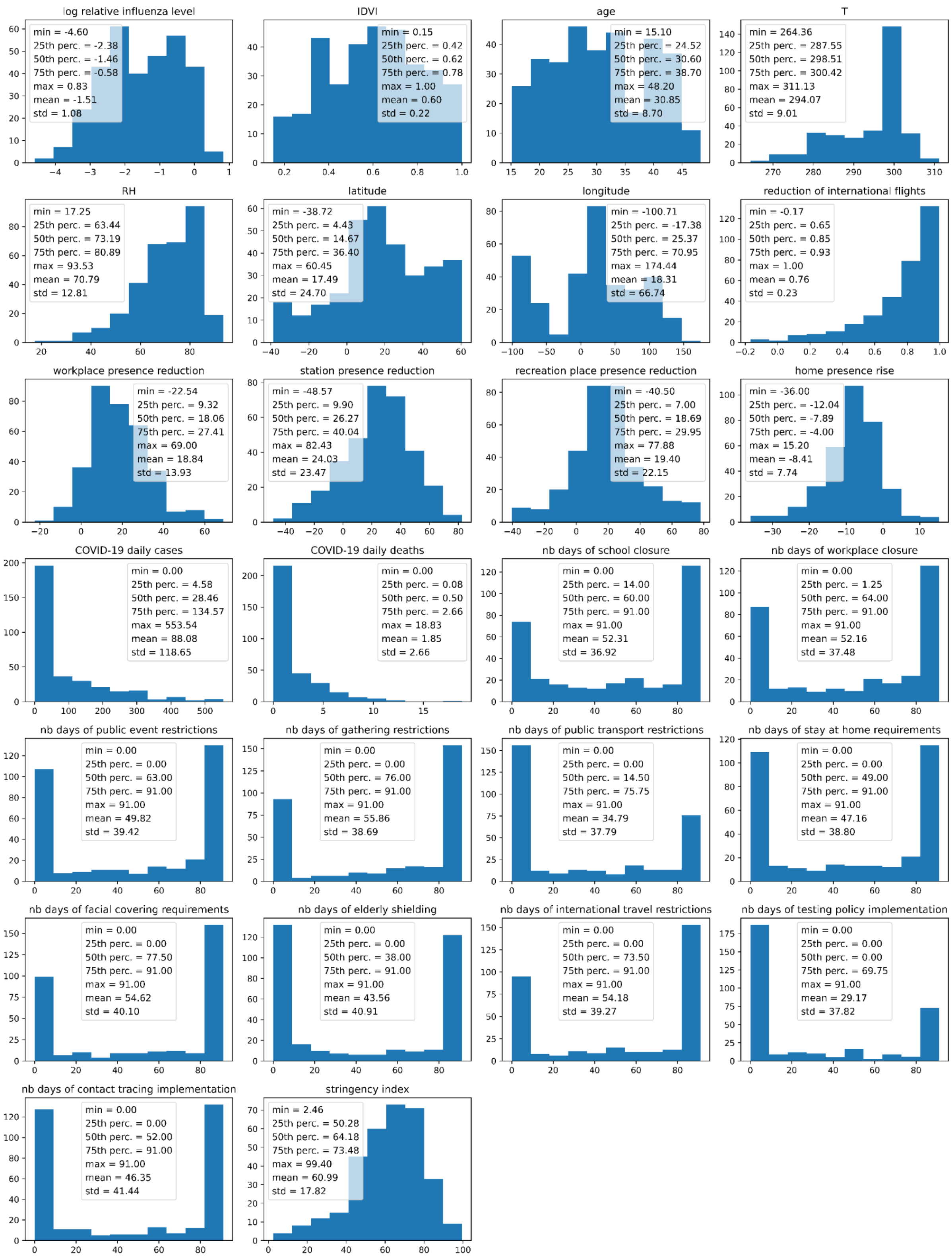


**Figure S1. Distributions of the 26 variables considered in the regression analysis, for all the 330 observations included in the study.** For each plot, summary values of the distributions are shown in the legend. We included here both the covariates considered in the main analysis and the covariates considered in the sensitivity analysis.

#### Algorithms for the regression analysis

Clustering and regression trees: We relied on the CART algorithm [2] to classify countries-trimesters based on a target variable, here the log relative influenza level. In a nutshell, observations are iteratively split in groups according to a covariate selected at each iteration so that the intra-group variance of the target variable is minimised. We controlled the structure of the tree by fixing the minimum number of observations in a terminal leaf (*minbucket*) and the regularisation parameter *cp* in the R package *rpart* [3]. The two parameters are optimised by cross-validation.

Cross-Validation for the hyperparameter tuning of the regression tree: The search for the optimal values of *minbucket* and *cp* is run over the following grid of parameters: $minbucket \in \{4 \leq m \leq 18 \vee m \in N\}$ and $cp \in \{0.001 * c \vee 0 \leq c \leq 20, c \in N\}$. For each point of the parameter space, 2000 trees were generated. Each tree is created on a random sample of 70% of the data and its prediction error (1-coefficient of determination) is calculated on the remaining 30% of the observations. Following Breiman' rule, the optimal tree is identified as the smallest tree that has mean prediction error less than the minimum error increased by its standard deviation. The simplest tree is identified by looking at the smallest number of splits on average, the largest *minbucket*, and the largest *cp*, in order. The optimal parameters identified were *cp=0.011, minbucket=4*.

Variable selection through permutation risk measures for covariate importance: The Variable Selection Using Random Forests (VSURF) algorithm has been exploited to identify the predictors associated with the reduction of influenza. This method evaluates the importance of each variable by measuring the prediction error increase when values of one variable at a time are permuted. This is a classical method used in the framework of Random Forests and, more in general, in machine learning algorithms. Also, some studies pointed out that permutation risk measures of variable importance are often more effective than alternative methods based on Sobol's indices or Shapley values [4].

The VSURF algorithm was run using the following parameters: *ntrees=8000, nfor.thres=100, nfor.interpr=100, nfor.pred=100* (and *mtry=6* by default).

### Additional results

#### Countries included in the analysis

There were 166 countries that contributed to FluNet during the period from 15 Dec 2014 to 12 Sep 2021. All FluNet records for these 166 countries were included in Figure 1A. Upon filtering based on the quality and extent of the FluNet records, 112 countries were included in the descriptive study (Figure 1B and Figure 2). For those countries, only the trimesters satisfying the inclusion criteria were included. Among the 112 countries, covariates were available only for 93 countries. This last group of countries was included in the regression analysis. The list of countries discarded at each step and included among the 93 countries is reported in Table S1.

| | countries in Fig. 1A (166) | | |
|---|---|---|---|
| | | countries in Fig. 1B and Fig. 2 (112) | |
| | | | countries for the regression analyses (93) |
| **Central and South America** | AIA, ATG, ABW, BHS, BRB, BLZ, VGB, CYM, CUB, DMA, DOM, GUF, GRD, GLP, MTQ, KNA, LCA, VCT, SUR, TTO, TCA, VEN | | ARG, BOL, BRA, CHL, COL, CRI, ECU, SLV, GTM, GUY, HTI, HND, JAM, MEX, NIC, PAN, PRY, PER, URY |
| **North America and Europe** | BEL, BMU, CZE, GRC, MLT, MNE, NLD, SVK, CHE | ALB, ISL, OWID_KOS, MKD | AUT, BLR, BIH, BGR, CAN, HRV, DNK, EST, FIN, FRA, DEU, HUN, IRL, ITA, LVA, LTU, LUX, NOR, POL, PRT, MDA, ROU, RUS, SRB, SVN, ESP, SWE, UKR, GBR, USA |
| **Africa** | BFA, CAF, TCD, MRT, MAR, RWA, SYC, SLE, SSD, TUN, ZWE | DZA, COD, ETH, GIN, MDG | AGO, CPV, CMR, COG, CIV, EGY, GMB, GHA, GNB, KEN, MLI, MUS, MOZ, NAM, NER, NGA, SEN, ZAF, SDN, TGO, UGA, TZA, ZMB |
| **Western, Central and South Asia** | BHR, CYP, KWT, MMR, SYR, TJK, TKM, ARE, UZB, YEM | ARM, AZE, BTN, IRN, MDV, TLS, PSE | AFG, BGD, KHM, GEO, IND, IDN, IRQ, ISR, JOR, KAZ, KGZ, LAO, LBN, MYS, NPL, OMN, PAK, PHL, QAT, SAU, SGP, LKA, THA, TUR, VNM |
| **Easthern Asia and Oceania** | FJI, PNG | CHN, PRK, NCL | AUS, JPN, MNG, NZL, KOR |

**Table S1. Countries included in the different steps of the study.** Countries are indicated with their 3-letter code, OWID_KOS is for Kosovo. Countries are grouped into five regions, aggregating different influenza transmission zones [5]: Central and South America (Temperate South America, Tropical South America and Central America and Caribbean), North America and Europe (North America, Northern Europe, South West Europe and Eastern Europe), Africa (Northern Africa, Western Africa, Middle Africa, Eastern Africa, Southern Africa), Western, Southern and Central Asia (Western Asia, Southern Asia, South-East Asia, Central Asia), Eastern Asia and Oceania (Eastern Asia, Oceania Melanesia Polynesia).

### Regression tree

<u>Additional details of the 5-group classification:</u> We provide in the following additional details on the 5-group repartition presented in Figure 4 of the main paper: the box plot of covariate values for observations in each group (Figure S2), and the list of countries-trimester belonging to each group (Table S2).

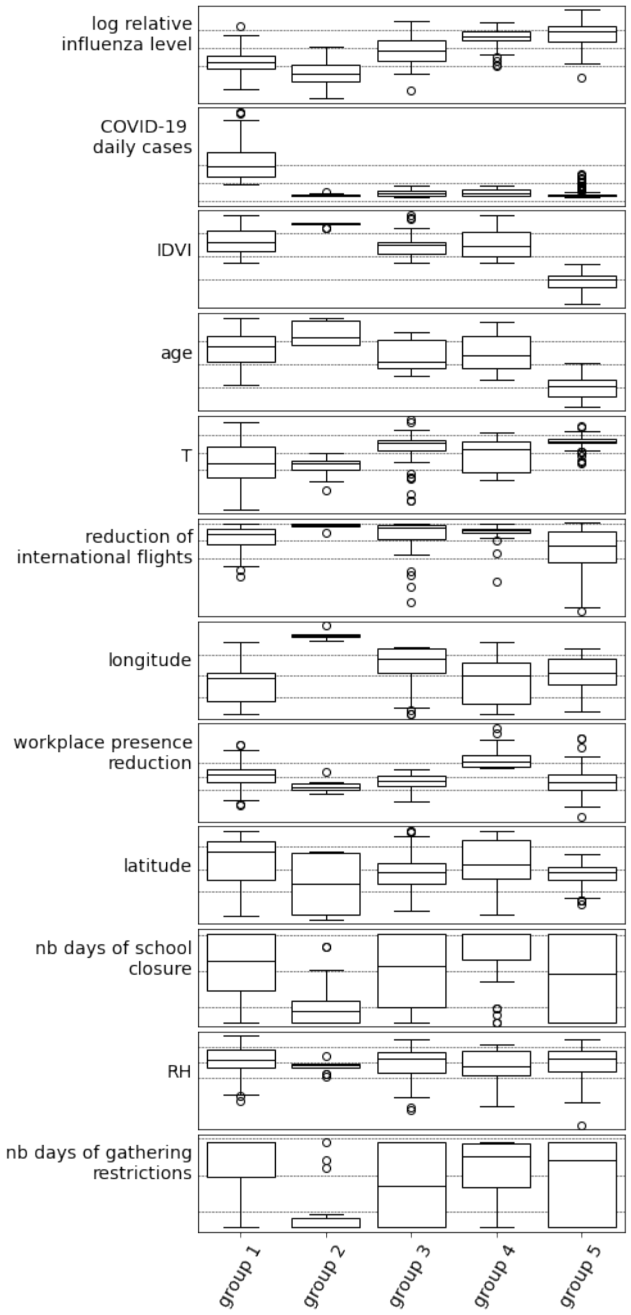


**Figure S2. Variable distributions for the five-group partitioning by means of the regression tree.** For each variable the boxplot shows the distribution in the group. Horizontal lines show median and quartiles of the whole dataset for comparison.

| | Spring 2020 | Summer 2020 | Autumn 2020 | Winter 2020-21 | Spring 2021 | Summer 2021 |
|---|---|---|---|---|---|---|
| **Group 1: leaves 1,2** | | | | | | |
| **Central and South America** | CHL | ARG, BRA, CHL, COL, CRI, PAN | ARG, CHL, COL, CRI | BRA, COL, CRI, MEX | ARG, BRA, CHL, COL, CRI, ECU, PAN, PER, PRY | ARG, BRA, CHL, COL, CRI, PAN, PRY, URY |
| **North America and Europe** | | | CAN, GBR, IRL, PRT, USA | AUT, BGR, BLR, CAN, DEU, DNK, ESP, EST, FRA, GBR, HRV, HUN, IRL, ITA, LUX, LVA, MDA, NOR, POL, PRT, ROU, RUS, SRB, SVN SWE, UKR, USA | AUT, BLR, CAN, DEU, DNK, EST, HRV, IRL, LTU, LUX, LVA, MDA, NOR, POL, ROU, SVN, SWE, UKR, USA | |
| **Africa** | | ZAF | | | GEO, JOR, LBN, MYS, OMN, QAT, TUR | ZAF |
| **Western, Southern and Central Asia** | QAT, SGP | QAT | ISR, JOR, LBN, OMN | GEO, ISR, JOR, LBN, QAT, TUR | | LKA, MYS, PHL, THA |
| **Eastern Asia and Oceania** | | | | | | |
| **Group 2: leaf 3** | | | | | | |
| **Central and South America** | | | | | | |
| **North America and Europe** | | | | | | |
| **Africa** | | | | | | |
| **Western, Southern and Central Asia** | | | | | | |
| **Eastern Asia and Oceania** | AUS, JPN | AUS, NZL | AUS, JPN, KOR | AUS, JPN, KOR | AUS, JPN, KOR | AUS |
| **Group 3: leaves 4,5,6,7** | | | | | | |
| **Central and South America** | BRA, PRY | PRY | MEX | | MEX | ECU, PER, SLV |
| **North America and Europe** | SWE | | NOR | FIN | RUS | |
| **Africa** | | MUS | ZAF | | ZAF | |
| **Western, Southern and Central Asia** | THA, VNM | IDN, LKA, MYS, THA, VNM | KAZ, MYS, QAT, SAU, SGP, THA, VNM | KAZ, LKA, SAU, SGP, THA, VNM | IDN, SAU, SGP, THA, VNM | QAT, SAU, SGP |
| **Eastern Asia and Oceania** | MNG | | | MNG | | |
| **Group 4: leaves 8,9** | | | | | | |
| **Central and South America** | ARG, COL, CRI, MEX, PAN, PER, SLV | SLV | | ECU | | |
| **North America and Europe** | AUT, CAN, DEU, DNK, EST, GBR, IRL, LVA, NOR, POL, ROU, RUS, SVN, UKR, USA | | | | GBR | |
| **Africa** | MUS, ZAF | | | | | |
| **Western, Southern and Central Asia** | IDN, LKA, MYS, OMN, SAU | SGP | LKA | IDN, KGZ, OMN | LKA, PHL | |
| **Eastern Asia and Oceania** | | | | | | |
| **Group 5: leaves 10,11,12,13,14** | | | | | | |
| **Central and South America** | BOL, GTM, HND, HTI, JAM | HND, HTI, NIC | BOL, HND, HTI, NIC | GTM, HND, HTI | BOL, GTM, HND, HTI, JAM | GTM, HND, HTI, JAM, NIC |
| **North America and Europe** | | | | | | |
| **Africa** | CIV, CMR, EGY, MLI, MOZ, TZA, ZMB | CIV, EGY, KEN, MLI, SEN, TZA, UGA, ZMB | CIV, CMR, EGY, GHA, KEN, NER, SEN, TGO, TZA, UGA, ZMB | CIV, CMR, EGY, GHA, KEN, NER, NGA, SEN, TGO, TZA, ZMB | CIV, CMR, EGY, GHA, KEN, MLI, NGA, SEN, TGO, TZA, UGA, ZMB | CIV, CMR, EGY, GHA, KEN, MLI, NGA, SEN, TGO, TZA, UGA, ZMB |
| **Western, Southern and Central Asia** | AFG, BGD, IND, KHM, LAO, NPL | BGD, KHM, LAO, NPL | AFG, BGD, IND, IRQ, KHM, LAO, NPL, PAK | AFG, IND, IRQ, KHM, LAO, NPL, PAK | AFG, BGD, IND, KHM, LAO, NPL, PAK | BGD, IND, KHM, LAO, NPL |
| **Eastern Asia and Oceania** | | | | | | |

**Table S2. Classification of countries-trimesters according to the high-level partitioning in five groups by means of the regression tree.** Countries are grouped into five regions, aggregating different influenza transmission zones [5]: Central and South America (Temperate South America, Tropical South America and Central America and Caribbean), North America and Europe (North America, Northern Europe, South West Europe and Eastern Europe), Africa (Northern Africa, Western Africa, Middle Africa, Eastern Africa, Southern Africa), Western, Southern and Central Asia (Western Asia, Southern Asia, South-East Asia, Central Asia), Eastern Asia and Oceania (Eastern Asia, Oceania Melanesia Polynesia).

<u>Full Regression Tree</u>: The regression tree selected using the algorithm had 14 terminal leaves and a coefficient of determination $R^2$=0.69. The leaves identified by the model were well defined (Figure S3), i.e. distinct from each other and characterised by homogeneous values of log relative influenza level - only for two of them the interquartile width of the observed log relative influenza level was greater than unity.

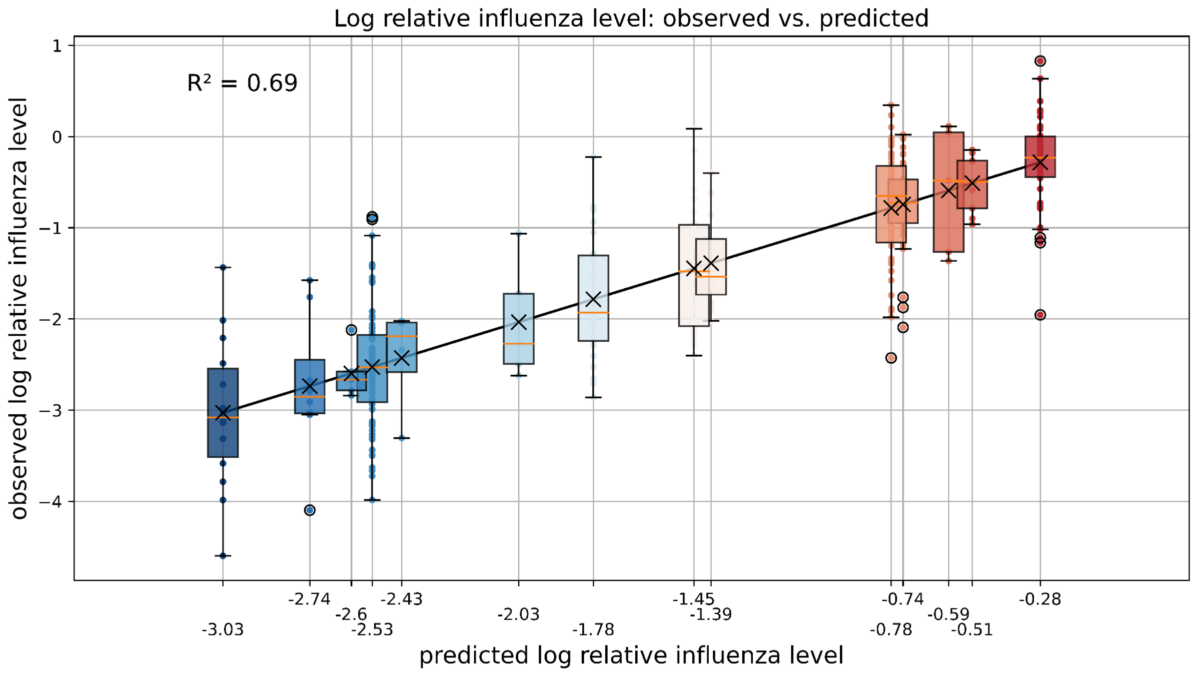


**Figure S3: Goodness of fit of the regression tree.** The 14 boxplots display the distributions of the log relative influenza level for countries-trimesters of the 14 leaves. The black crosses identify the mean values of the distributions (also shown through the color scale) that correspond to the predicted log relative influenza level. $R^2$ is the coefficient of determination.

The tree is shown in Figure S4. The first four splits are done according to IDVI, COVID-19 daily cases, longitude and workplace presence reduction as discussed in the main paper. The other variables are used for a finer partition in smaller groups. The classification of countries-trimesters in leaves is reported in the supplementary data [6].

Group 1 (109 observations) is split into leaves 1 and 2. In leaf 1, lower temperatures relative to leaf 2 are associated with a greater reduction in influenza. Leaf 1 consists largely of temperate countries in Europe, North and South America, during the 2020-2021 influenza season, which had a greater influenza reduction. Leaf 2 includes a more limited number of observations (26, compared with the 83 in leaf 1) from countries of tropical and subtropical areas with IDVI>0.54, e.g. Panama, Costa Rica, Colombia, Malaysia. Influenza reduction for these countries was less strong compared with leaf 1, but still substantial if compared with low-IDVI tropical countries, classified in group 5.

Group 2 is formed by a single leaf with well defined properties detailed in the main paper.

The 45 observations of the group 3 are distributed in four leaves including a few countries-trimesters each. Similarly to the split within group 1, a first split based on temperature separates countries with higher temperature and higher log relative influenza level (Saudi Arabia and Qatar) from countries with lower temperature and lower log relative influenza level. This second branch splits based on the reduction of international flights. On the left side of the split there is a group of 8 countries-trimesters where low values of log relative influenza level were associated with lower-than-average reduction of international flights, reduction of workplace presence and number of days with gathering restrictions. This group was characterised by a number of days of school closure higher than average. The right side of the split has two leaves that are discussed in the main paper, i.e. leaf 5 including mainly Singapore and leaf 6 including mainly other Southeast Asia countries.

Group 4 consists of 39 observations. This includes mainly countries during spring 2020 that are grouped in leaf 9 (34 out of 39 observations). Five observations are separated by the other because they have a more limited number of days with school closure and a lower log relative influenza level. This is a heterogeneous set of countries, mainly between winter 2021 and spring 2021.

Group 5 contains a significant proportion of all observations (123 out of 330) that are separated into five leaves (leaves 10, 11, 12, 13 and 14). Interestingly, the five leaves show a clear trend with increasing log relative influenza level that is, in general, associated with a decrease in four of the five COVID-19 response variables - COVID-19 daily cases, reduction of international flights, reduction of workplace presence, and number of days with school closure. Limitations on gatherings remain moderate for all five leaves. Two splits are based on the reduction of international flights, between leaf 10 and leaves 11 and 12, and between leaf 13 and 14. For these two splits a greater reduction of international flights is associated with a lower influenza log ratio. Finally, leaves 11 and 12 differ in relative humidity. The two leaves contain almost the same set of countries for different trimesters - e.g. Guatemala, Honduras, India, Nepal, and Zambia. These are tropical countries characterised by a dry and a rainy season throughout the year, where influenza usually peaks twice a year with the main peak during the rainy season [7–10]. Our analysis shows that for rainy seasons the reduction of influenza was smaller.

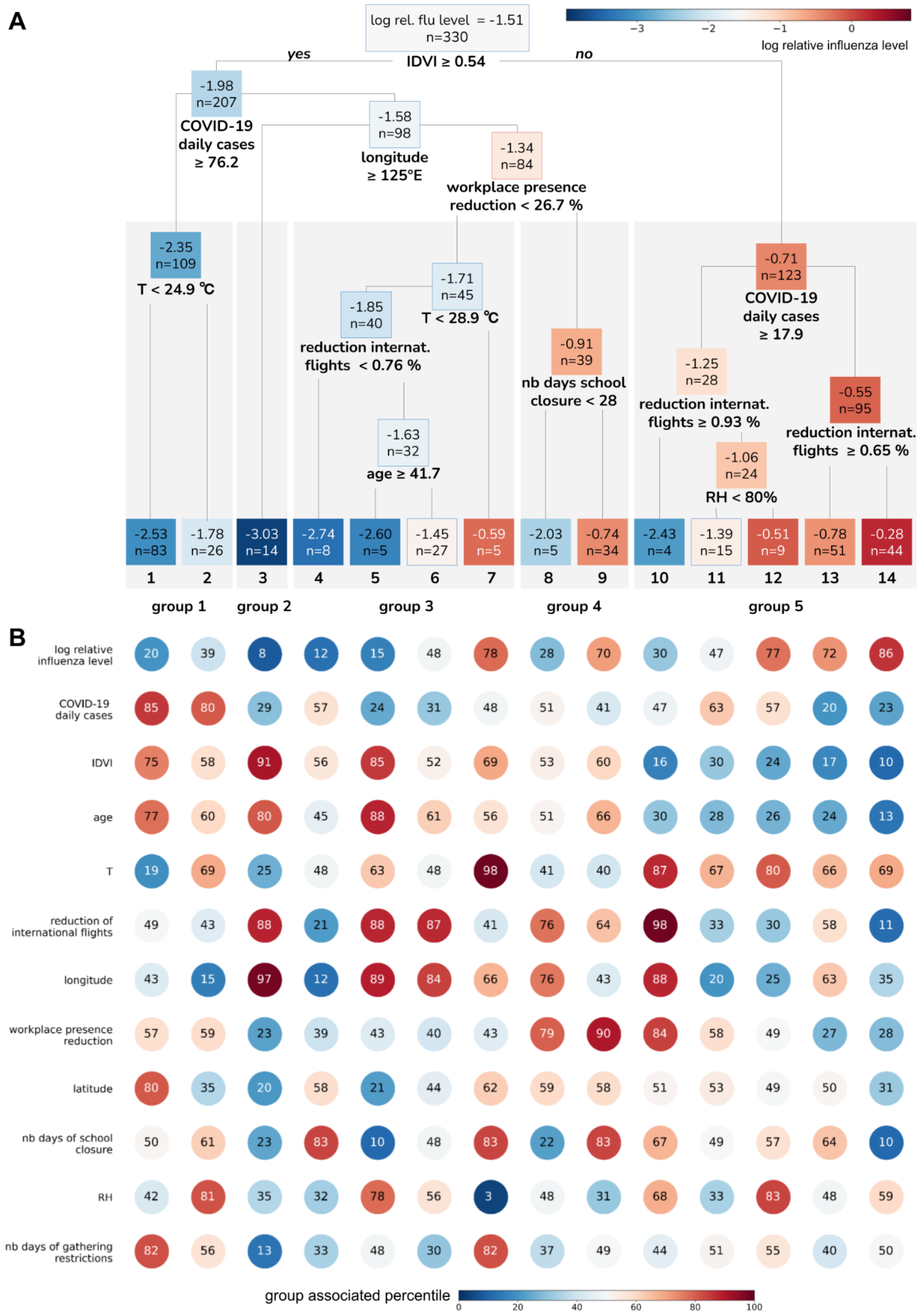


| | 1 | 2 | 3 | 4 | 5 | 6 | 7 | 8 | 9 | 10 | 11 | 12 | 13 | 14 |
|---|---|---|---|---|---|---|---|---|---|---|---|---|---|---|
| Leaf mean | -2.53 | -1.78 | -3.03 | -2.74 | -2.60 | -1.45 | -0.59 | -2.03 | -0.74 | -2.43 | -1.39 | -0.51 | -0.78 | -0.28 |
| n | 83 | 26 | 14 | 8 | 5 | 27 | 5 | 5 | 34 | 4 | 15 | 9 | 51 | 44 |
| log relative influenza level | 20 | 39 | 8 | 12 | 15 | 48 | 78 | 28 | 70 | 30 | 47 | 77 | 72 | 86 |
| COVID-19 daily cases | 85 | 80 | 29 | 57 | 24 | 31 | 48 | 51 | 41 | 47 | 63 | 57 | 20 | 23 |
| IDVI | 75 | 58 | 91 | 56 | 85 | 52 | 69 | 53 | 60 | 16 | 30 | 24 | 17 | 10 |
| age | 77 | 60 | 80 | 45 | 88 | 61 | 56 | 51 | 66 | 30 | 28 | 26 | 24 | 13 |
| T | 19 | 69 | 25 | 48 | 63 | 48 | 98 | 41 | 40 | 87 | 67 | 80 | 66 | 69 |
| reduction of international flights | 49 | 43 | 88 | 21 | 88 | 87 | 41 | 76 | 64 | 98 | 33 | 30 | 58 | 11 |
| longitude | 43 | 15 | 97 | 12 | 89 | 84 | 66 | 76 | 43 | 88 | 20 | 25 | 63 | 35 |
| workplace presence reduction | 57 | 59 | 23 | 39 | 43 | 40 | 43 | 79 | 90 | 84 | 58 | 49 | 27 | 28 |
| latitude | 80 | 35 | 20 | 58 | 21 | 44 | 62 | 59 | 58 | 51 | 53 | 49 | 50 | 31 |
| nb days of school closure | 50 | 61 | 23 | 83 | 10 | 48 | 83 | 22 | 83 | 67 | 49 | 57 | 64 | 10 |
| RH | 42 | 81 | 35 | 32 | 78 | 56 | 3 | 48 | 31 | 68 | 33 | 83 | 48 | 59 |
| nb days of gathering restrictions | 82 | 56 | 13 | 33 | 48 | 30 | 82 | 37 | 49 | 44 | 51 | 55 | 40 | 50 |

**Figure S4: Full regression tree. A** Regression tree obtained with the variables selected in Figure 3. For each node the average log relative influenza level and the number of observations are reported (the former is also indicated with a colour scale). **B** Properties of each leaf. For each covariate the percentile of the whole dataset distribution the median of the group corresponds to is indicated with the colour scale and reported in the bubble.

### Robustness checks and sensitivity analyses

#### Robustness of the variable selection procedure

The variable selection procedure is a stochastic algorithm that may lead to different results when repeated. Therefore, variable importance was estimated by averaging over 100 stochastic realisations. To be sure results were stable we repeated the procedure 20 times. Each time the same 11 predictors are selected as the important covariates for predicting the log relative influenza ratios.

#### Robustness of the tree structure optimization

The regression tree was regularised by two parameters (*minbucket* and *cp*) optimised with cross-validation through the stochastic procedure discussed in the Additional methods section of the supplementary information. To be sure that the procedure converges to a stable result we repeated it 10 times. The regularisation parameters selected each time were very similar and led to the construction of trees almost identical to the tree described in the Results section and in the supplementary information. In particular, the classification of countries-trimesters in the five high-level groups was robust.

#### Robustness of the tree under small perturbations of the dataset

We assessed the robustness of the tree under small perturbations in the dataset. We built ten regression trees on a random subsample of 314 observations (~95% of the total) keeping the same 11 predictors and hyperparameters. The ten resulting trees (i.e. the perturbed trees) were compared with the tree built from the entire dataset (i.e. the reference tree) using the Adjusted Rand Index (ARI) [11]. Specifically, we compared the classification in 5 groups to assess the robustness of the 5-group repartition discussed in the main paper. The average score value for the ten comparisons is 0.86, indicating good agreement between the perturbed and reference trees.

#### Sensitivity of the variable selection under changes on the assumption made

We tested whether the predictors of the log relative influenza level changed with the choices made throughout the analysis. The creation of the dataset of observations is based on two main assumptions: (i) *k=0.5* positive cases had been added to each country-trimester in order to remove zero counts of influenza cases, and (ii) a threshold *s=130* for the minimum number of tests processed per quarter was set to discard countries-trimesters with poor data. In addition, some choices were made when defining the covariates. We tested the robustness of our results to all these choices by analysing the following alternative models (the baseline model is here referred as *base 0* model):

- *base 1*: k=1 (instead of 0.5), s=130, same covariates of the base 0 model;
- *base 2*: k=0.5, s=26 (instead of 130), same covariates of the base 0 model;
- *base 3*: k=0.5, s=260 (instead of 130), same covariates of the base 0 model;
- *Cov 1*: k=0.5, s=130, COVID-19 daily deaths is used in alternative to COVID-19 daily cases to quantify the intensity of the COVID-19 epidemic;
- *Mob 1:* k=0.5, s=130, the public transport station presence reduction is tested in alternative to workplace presence reduction to capture changes of social activity;
- *Mob 2:* k=0.5, s=130, the recreation place presence reduction is tested in alternative to workplace presence reduction to capture changes of social activity;

- *Mob 3:* k=0.5, s=130, the increase in home presence is tested in alternative to workplace presence reduction to capture changes of social activity;
- *No Age:* k=0.5, s=130, the variables age is removed among the set of covariates to be included in the regression;
- *No IDVI:* k=0.5, s=130, the variables IDVI is removed among the set of covariates to be included in the regression;
- *Str. Idx:* k=0.5, s=130, all variables associated with NPIs are replaced by the stringency index.

For all the alternative models the selected sets of important factors are highly similar (Table S3): impact of COVID-19, international mobility, workplace presence reduction (or the alternative proxy of social activity considered), IDVI, age, temperature and longitude always result significant, while latitude and RH are discarded only once and twice respectively. All proxies of social activity tested were classified as important. The stringency index when included was not selected, indicating that the aggregate information it carries is not important in explaining the influenza reduction. This is consistent with the fact that only 2 out of the 11 governamental response variables were selected as important.

We used the ARI similarity index to compare the sensitivity trees and the baseline tree up to the five-group repartitions. Models Mob 1, Mob 2, Mob 3, No Age, Str. Idx. led to an excellent recovery of the baseline tree (ARI > 0.9). The 5-group repartition showed the same behaviour as in the baseline model. The tree obtained with Base 1 was in good agreement with the baseline tree. Interestingly, removing IDVI from the set of covariates led to only a moderate recovery of the baseline tree (ARI= 0.69), despite IDVI and Age being highly correlated. Age is not able to fully compensate for IDVI in creating the 5 group repartition discussed in the main paper. This is consistent with the fact that both covariates were found to be important by the VSURF procedure.

The comparison of trees obtained with base 2, base 3 and Cov 1 and the baseline tree up to the five-groups repartition led to lower similarity values (ARI between 0.53 and 0.66). Still, we found that the four trees shared in large part the same behaviour. In particular, the first split was the same (i.e. based on IDVI with the same threshold value), meaning that all partitions had the distinction between high IDVI, low influenza countries and low IDVI, high influenza ones. Also, the 2020 spring trimesters of temperate countries (comprising the bulk of group 4) remained in great part grouped together and separated from the rest. In all three sensitivity trees, countries of group 2 of the baseline tree (zero-covid countries) were grouped together and included in a larger group together with countries of group 1 of the baseline tree.

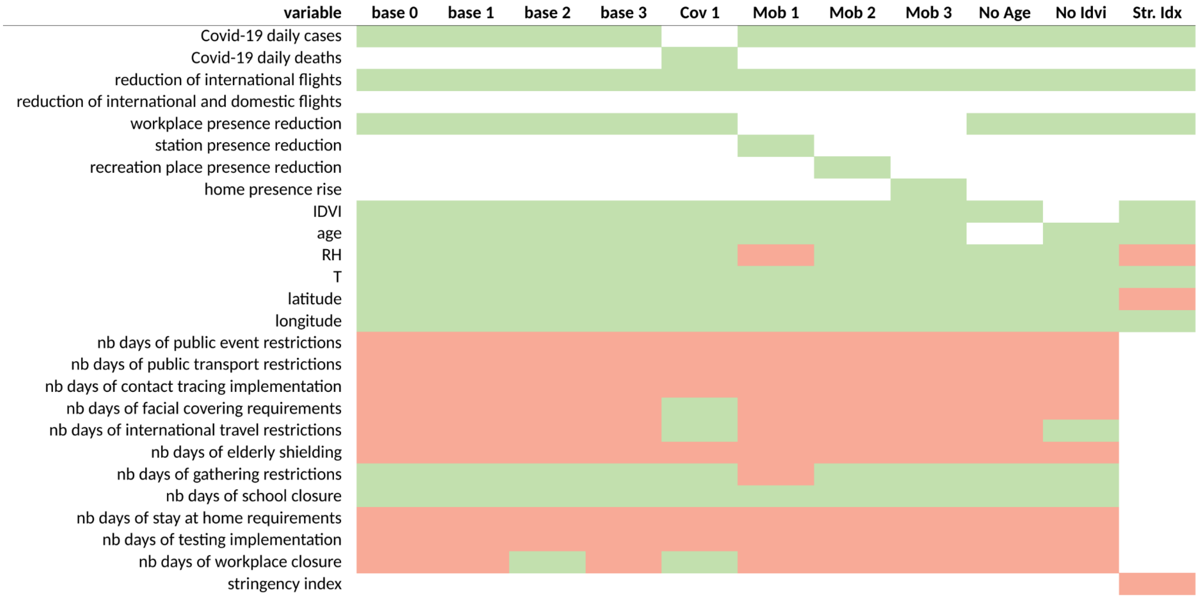


| variable | base 0 | base 1 | base 2 | base 3 | Cov 1 | Mob 1 | Mob 2 | Mob 3 | No Age | No Idvi | Str. Idx |
|---|---|---|---|---|---|---|---|---|---|---|---|
| **k** | 0.5 | 1 | 0.5 | 0.5 | 0.5 | 0.5 | 0.5 | 0.5 | 0.5 | 0.5 | 0.5 |
| **s** | 130 | 130 | 26 | 260 | 130 | 130 | 130 | 130 | 130 | 130 | 130 |
| **nb observations** | 330 | 335 | 390 | 279 | 321 | 330 | 330 | 330 | 330 | 330 | 330 |
| **nb tested predictors** | 20 | 20 | 20 | 20 | 20 | 20 | 20 | 20 | 19 | 19 | 10 |
| **nb selected predictors** | 11 | 11 | 12 | 11 | 14 | 9 | 11 | 11 | 10 | 11 | 7 |
| **Similarity index (ARI)** | – | 0.82 | 0.61 | 0.53 | 0.66 | 0.94 | 0.95 | 0.92 | 1.00 | 0.69 | 1.00 |

**Table S3. Predictors selected for 11 alternative models.** Each model includes only variables associated with a colored cell, green is for the selected variables, red for the rejected ones. Additional information about the parameters *k,s* used for the definition of the observation set is provided. Also, the similarity index is reported: it measures the similarity of the five-group classifications made by each alternative model compared with the reference model *(base 0*).
